# Impact of the COVID-19 pandemic on the mental health and wellbeing of parents with young children: a qualitative interview study

**DOI:** 10.1101/2021.05.13.21256805

**Authors:** Jo Dawes, Tom May, Alison McKinlay, Daisy Fancourt, Alexandra Burton

**Author notes:** Corresponding author: Alexandra Burton.

## Abstract

**Background:** Parents have faced unique challenges during the coronavirus disease 2019 (COVID-19) pandemic, including mobility constraints, isolation measures, working from home, and the closure of schools and childcare facilities. There is presently a lack of in-depth qualitative research exploring how these changes have affected parents’ mental health and wellbeing.

**Methods:** Semi-structured qualitative interviews with 29 parents of young children. Interviews were analysed using reflexive thematic analysis.

**Results:** We identified five superordinate themes affecting participant mental health and wellbeing: 1) navigation of multiple responsibilities and change inside the home; 2) disruption to home life; 3) changes to usual support networks; 4) changes in personal relationships; and 5) use of coping strategies. Participants described stress and exhaustion from navigating multiple pressures and conflicting responsibilities with home, schooling, and work, without their usual support networks and in the context of disrupted routines. Family roles and relationships were sometimes tested, however, many parents identified coping strategies that protected their wellbeing including access to outdoor space, spending time away from family, and avoiding conflict and pandemic-related media coverage.

**Conclusions:** Employers must be cognisant of the challenges that the pandemic has placed on parents, particularly women and lone parents. Flexible working arrangements and support might therefore relieve stress and increase productivity. Coping strategies identified by parents in this study could be harnessed and encouraged by employers and policymakers to promote positive wellbeing during times of stress throughout the pandemic and beyond.

## Background

In response to the coronavirus disease 2019 (COVID-19) pandemic, restrictions of varying stringency were implemented to reduce the transmission of the virus. In the United Kingdom (UK), this included mobility constraints, self-isolation and the closure of non-essential businesses and workplaces, hospitality, and educational settings. Whilst necessary to control the virus, such measures disrupted the lives of various groups of the population, with many reporting psychological distress, social isolation, and economic hardship (1-3).

The impact of the COVID-19 pandemic presented specific challenges to parents of young children and their families. In the UK, the swift closure of education and childcare settings led to many parents suddenly being responsible for day-care and home-schooling their children alongside existing work and household responsibilities. Research suggests that the consequential increase in childcare responsibilities at home (4) alongside reduced contact with social networks led to increased parental stress (5, 6). In particular, parents who perceived themselves to be less capable in home-schooling their children (whether due to a lack of resources, support, or other family commitments) experienced significant psychological distress (7, 8) as did families experiencing economic hardship (9).

In addition to navigating their own feelings of uncertainty in the early stages of the pandemic, parents also managed their children’s physical, psychological, and social wellbeing. Evidence suggests children experienced boredom, worry, frustration, loneliness and irritability (8), all of which affected parental distress and home-schooling during lockdowns (10, 11). This led third sector services and mental health organisations in the UK to express concern about psychological harm amongst parents during the pandemic (12). Concerns were heightened amongst certain groups such as parents of children with neurodevelopmental disorders who reported stress and challenges associated with the closure of services that usually provided specialised social and mental health support to families (8).

As well as the closure of schools and nurseries, the pandemic also had a major impact on the nature and structure of family life with reduced access to informal support networks such as family and friends. Some parents were able to adopt reduced or flexible working hours to accommodate the increased demands to cope with childcare (13). However, it is not clear how widespread these initiatives were for parents of young children. Further, there is limited evidence about the specific issues faced by parents, such as *how* these issues affected mental health and what actions parents took to support positive wellbeing. Answering these questions is important, both as the pandemic continues, to provide more targeted support and advice to parents, as well as in planning for future pandemics. Accordingly, there is a need for research that can provide in-depth data on the experiences of parents during the pandemic. The aim of this study was therefore to understand how the COVID-19 pandemic and associated social distancing restrictions affected parent mental health and well-being.

## Methods

We conducted a semi-structured qualitative interview study with parents of young children. The study formed part of the COVID-19 Social Study (14, 15). Ethical approval was granted by the University College London research ethics committee (Project ID: 14895/005).

### Sample and recruitment

We recruited participants via social media, personal contacts, the UK Research and Innovation (UKRI) funded MARCH Network, and the COVID-19 Social Study newsletter and website. Participants were eligible to take part in the study if they were aged 18 or over, living in the UK, and had at least one child aged 12 years or under. From those that responded to a researcher via email, demographic information was collected to allow for purposive sampling and to maximise representation of diversity across sex, age, ethnicity, living arrangements, age of children, and number of children. All parents who were invited to participate were emailed a participant information sheet and given an opportunity to ask questions either via email or by telephone/video call. All those who agreed to participate electronically signed an informed consent form prior to the interview. Recruitment to the study continued until the research team had spoken to individuals from a range of demographic backgrounds and until no new major themes were identified by the researchers.

### Data collection

Interviews were undertaken between June and November 2020 and lasted between 39 and 74 minutes (mean: 55 minutes). Interviews followed a semi-structured format using a Topic Guide of questions (please see additional file 1) informed by existing theories of behaviour change (16), social networks and mental health (17). Participants were interviewed remotely in their homes, having been advised to find a quiet space on their own to maintain confidentiality. Interviews were conducted via telephone or video call depending on participant preference. Interviewers were female and male researchers with experience of qualitative methods (JD is a research fellow in public health and healthcare clinician; AB, AR and AM are postdoctoral research fellows in social science and behavioural health; HA is a pre-doctoral researcher and trainee medic). Participants were not known to the research team prior to interview. Introductory information about the research project and the research team were explained verbally before the interview commenced. All interviews were audio-recorded and transcribed verbatim by an external transcription company. All participants who completed an interview were emailed an online £10 shopping voucher to thank them for their time.

### Data analysis

We used a reflexive thematic analysis approach (18) to analyse the data. Data were managed using NVivo 12 Pro software. JD was the primary data coder who read and coded all transcripts line-by-line. An initial coding framework was developed a priori based on key concepts from the interview topic guide. The framework was then applied to the transcripts and updated with new codes in response to content encountered in the transcripts. To ensure the consistency of the coding approach in identifying pertinent topics, a second researcher (AM) double coded 10% of the transcripts and coding was then compared and discussed between the researchers. On completion of coding, preliminary themes were generated by JD. The coding framework and subsequent themes were presented to the research team at weekly meetings for discussion and feedback. Themes were refined based on this feedback, finalised, and reported with supporting quotes from participants.

## Results

We recruited 29 parents of young children. Participants had on average two children (range 1-4 children) with a mean child age of 5 years (range 10 months to 12 years old). Demographics of the participant population are described in Table 1. Participants were aged 28-58 years (mean age 40), predominantly female (68.9%), white British (62.1%) with the majority (89.7%) educated to at least undergraduate/professional level. 86.3% described themselves as married, in a civil partnership or living with a partner. 75.9% lived with both their partner and children. 13.8% lived only with their children.

**Table 1.**
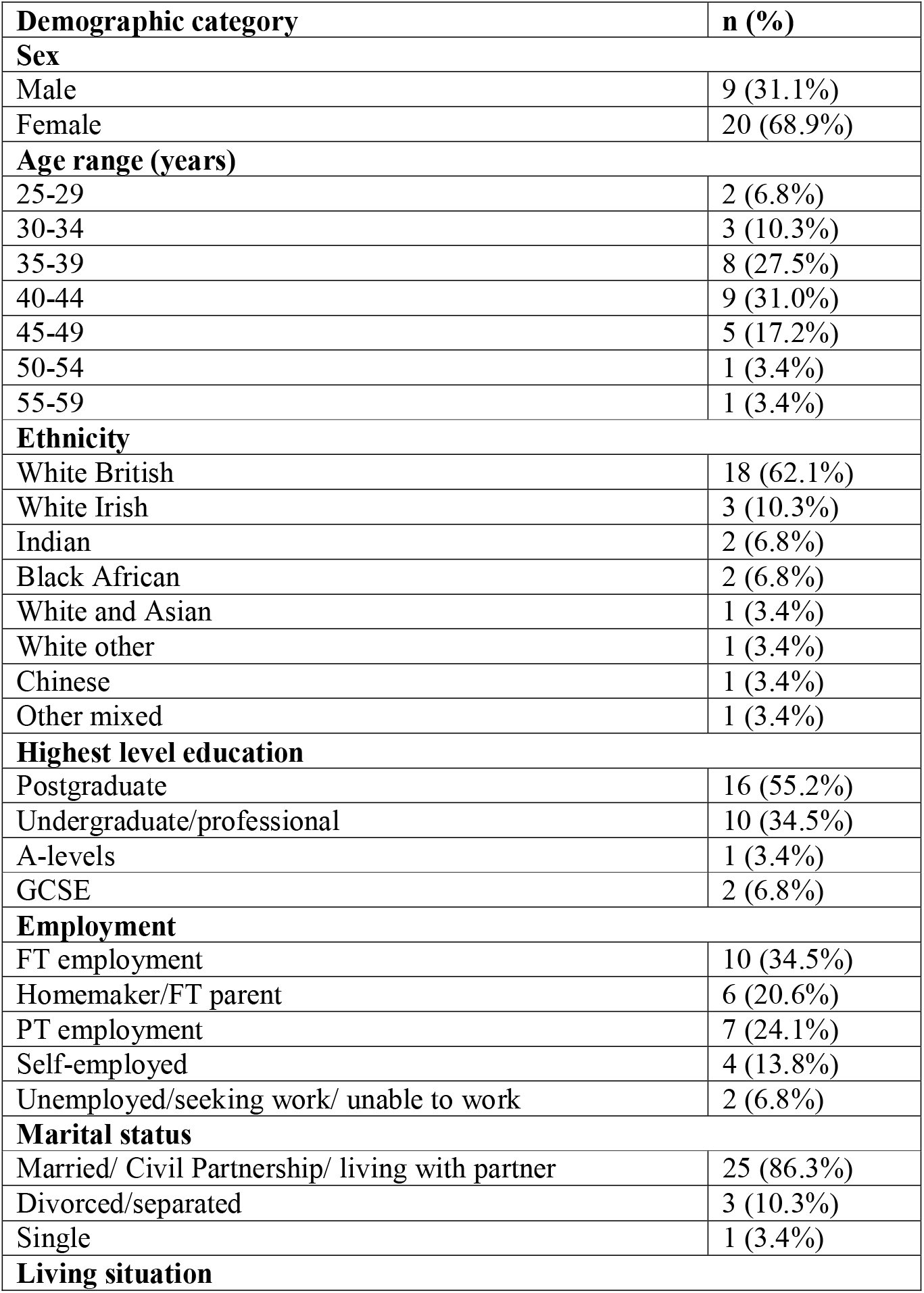
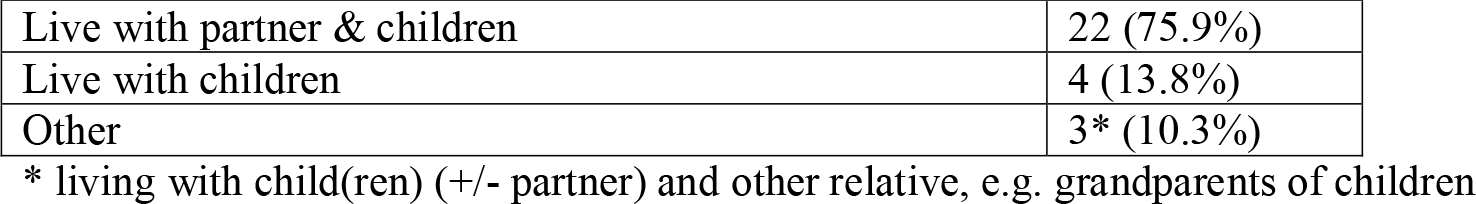
Participant demographics.

| **Demographic category** | **n (%)** |
| --- | --- |
| **Sex** |  |
| Male | 9 (31.1%) |
| Female | 20 (68.9%) |
| **Age range (years)** |  |
| 25-29 | 2 (6.8%) |
| 30-34 | 3 (10.3%) |
| 35-39 | 8 (27.5%) |
| 40-44 | 9 (31.0%) |
| 45-49 | 5 (17.2%) |
| 50-54 | 1 (3.4%) |
| 55-59 | 1 (3.4%) |
| **Ethnicity** |  |
| White British | 18 (62.1%) |
| White Irish | 3 (10.3%) |
| Indian | 2 (6.8%) |
| Black African | 2 (6.8%) |
| White and Asian | 1 (3.4%) |
| White other | 1 (3.4%) |
| Chinese | 1 (3.4%) |
| Other mixed | 1 (3.4%) |
| **Highest level education** |  |
| Postgraduate | 16 (55.2%) |
| Undergraduate/professional | 10 (34.5%) |
| A-levels | 1 (3.4%) |
| GCSE | 2 (6.8%) |
| **Employment** |  |
| FT employment | 10 (34.5%) |
| Homemaker/FT parent | 6 (20.6%) |
| PT employment | 7 (24.1%) |
| Self-employed | 4 (13.8%) |
| Unemployed/seeking work/ unable to work | 2 (6.8%) |
| **Marital status** |  |
| Married/ Civil Partnership/ living with partner | 25 (86.3%) |
| Divorced/separated | 3 (10.3%) |
| Single | 1 (3.4%) |
| **Living situation** |  |

| Demographic category | n (%) |
| --- | --- |
| Live with partner & children | 22 (75.9%) |
| Live with children | 4 (13.8%) |
| Other | 3* (10.3%) |

\* living with child(ren) (+/- partner) and other relative, e.g. grandparents of children

Five themes were identified (Table 2) relating to the impact of the COVID-19 pandemic and associated restrictions on mental health and wellbeing including: i) navigation of multiple responsibilities and change inside the home; ii) disruption to home life; iii) changes to usual support networks; iv) changes to personal relationships; v) use of coping strategies.

**Table 2.**
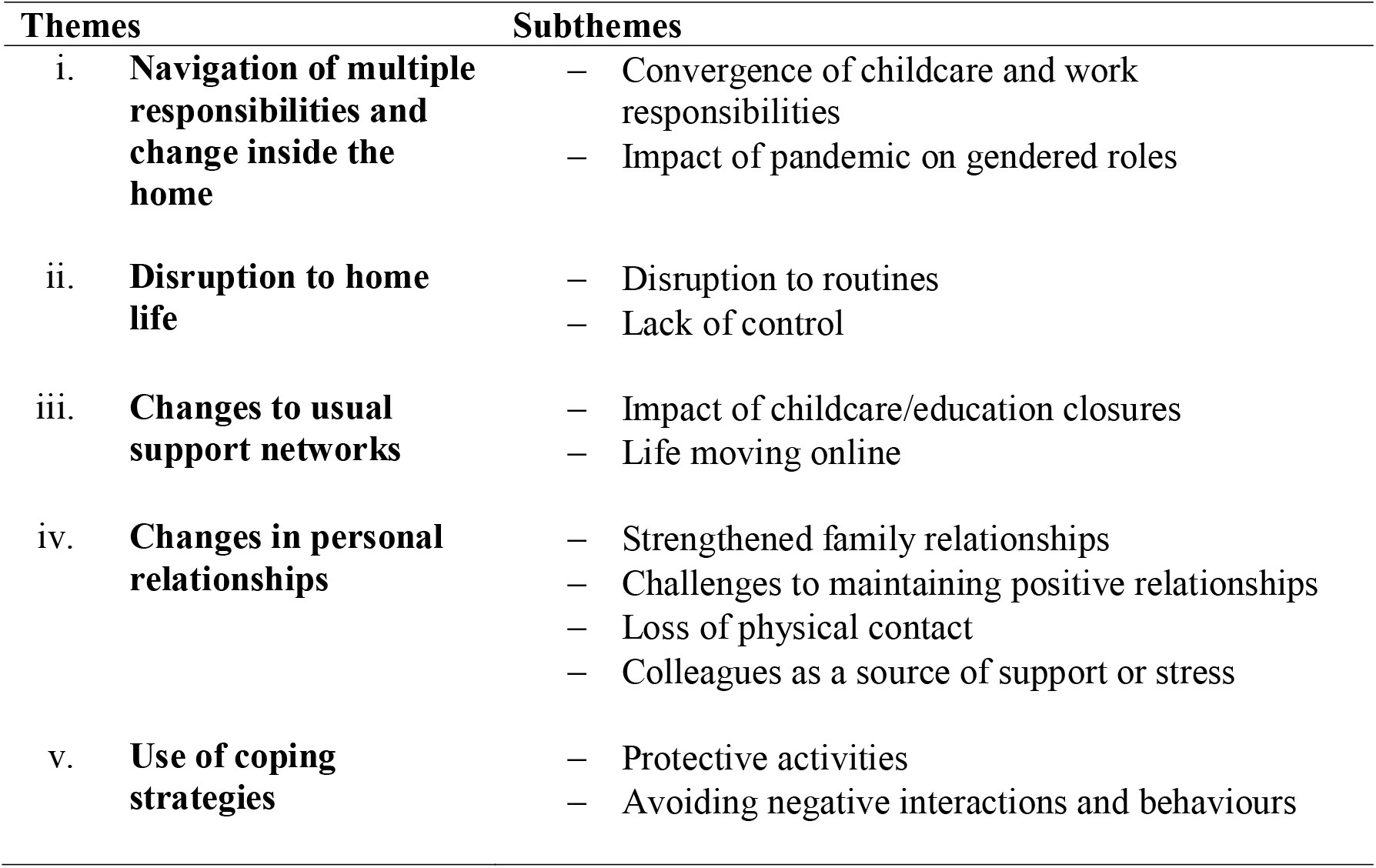
Factors influencing the mental health and wellbeing of parents of young children during the COVID-19 pandemic.

| Themes | Subthemes |
| --- | --- |
| i. **Navigation of multiple responsibilities and change inside the home** | – Convergence of childcare and work responsibilities<br>– Impact of pandemic on gendered roles |
| ii. **Disruption to home life** | – Disruption to routines<br>– Lack of control |
| iii. **Changes to usual support networks** | – Impact of childcare/education closures<br>– Life moving online |
| iv. **Changes in personal relationships** | – Strengthened family relationships<br>– Challenges to maintaining positive relationships<br>– Loss of physical contact<br>– Colleagues as a source of support or stress |
| v. **Use of coping strategies** | – Protective activities<br>– Avoiding negative interactions and behaviours |

### Navigation of multiple responsibilities and change inside the home

#### Convergence of childcare and work responsibilities

Participants commonly described challenges in managing multiple responsibilities within the home, many of which had previously occurred within separate and distinct environments. This included caring responsibilities, their children’s education and developmental needs, work, and day-to-day household responsibilities.

> “It just feels like I’ve got about 500 balls in the air at the same time and keeping them all in the air is hard work.” (ID384_lone_mum)

Some mothers, but mostly fathers described difficulties in not being able to fulfil their work responsibilities and care for young children concurrently.

> “Things became pretty unsatisfactory. From a work life balance point of view, as I said, not quite 24/7 childcare but it was definitely a lot more than I was used to, in pretty difficult circumstances and then, feeling like I was not really managing to get even half my work done.” (ID385_couple_dad)

Having to choose between prioritising work and parenting young children weighed heavily on the conscience of many participants. This often resulted in feelings of guilt, stress, and a sense that they were unable to fully attend to either role.

> “I guess there was a sense of guilt… especially when the children were at home when they were, not neglected as such, but balancing trying to work and interact with them was tricky.” (ID393_couple_dad)

When home schooling was added to competing work pressures, this sense of guilt was heightened. The pressure to prioritise work over their children’s education was particularly difficult for lone parents.

> “And my ability to do any of the…even supervising their education at home was very, very limited. And from a home perspective, my main concerns were about feeling guilty that I wasn’t able to give the children what other parents seemed to be giving their children in terms of support around their home education and just time with me.” (ID384_lone_mum)

One mother described how home schooling had challenged her identity and confidence as a mother, resulting in feelings of doubt and lowered self-esteem.

> “The one thing I thought I was good at, was that I was a good mum. And then because of the home schooling and how that made me feel, I started to feel like I wasn’t a good mum, so that made me feel like I wasn’t good at anything.” (ID386_lone_mum)

The move to home working and childcare was particularly challenging for parents with very young children, with many participants finding it difficult to explain the new arrangements to their children and manage their expectations.

> “It was a real upheaval having my son out of nursery and he was massively confused by why dad was at home, but he couldn’t play with him. I have a photo of him sitting outside the spare bedroom door on a cushion waiting for my husband to get off a conference call.” (ID394_couple_mum)

#### Impact of pandemic on gendered roles

Mothers, more so than fathers, tended to highlight how pandemic-related disruptions impacted their ability to manage usual household tasks (such as tidying away children’s toys, meal preparation for the household, washing up, cleaning, and buying food) alongside making time for work and/or home schooling.

> “I mean, normally, we do the cleaning and the tidying when the kids aren’t here, because let’s face it, it is easier. There’s that saying that trying to clean when the kids are at home, is like trying to brush their teeth while eating Oreos.” (ID301_lone_mum)

A common source of frustration for mothers was a perception that they were being made to fulfil traditional gender roles, especially when their children became aware of this division of labour. For some, conflict arose when there was a perceived imbalance between parental and work responsibilities.

> “It bothered me that the children saw us in such a traditional gendered split, with my husband doing a job for his boss, and me running the household. My husband would come downstairs for lunch and afterwards “go back to work.” My son asked a few times why Daddy was working, and why I didn’t work. I’d explain that I did work, that my work at the moment was to…take care of the house and family day-to-day and that I did have a job with a boss just like Daddy, that I just wasn’t doing right now…I definitely felt…that real clash of, you’re upstairs at work doing important things, and I’m down here covered in Play-Doh and snot.” (ID394_couple_mum)

For many mothers, the pandemic coincided with a transitional phase for the family, for example, the adoption of a child, finishing maternity leave, or children starting school. Coping with alterations to a family dynamic and the concurrent changes imposed by the pandemic was described as difficult. One mother spoke about the plans she had made for when her son started school, which were thwarted by the pandemic.

> “I’ve felt very resentful, honestly. This was supposed to be my year where I was allowed to focus a bit on me again. I have been the main childcare person in the family. With [name] going to school it was like okay, I was going to be allowed a bit more structure and I could rule my time again a bit more. So, I was resentful, I suppose, that was off the cards.” (ID383_couple_mum)

### Disruption to home life

#### Disruption to routines

Participants frequently described how difficult they found disruption to their usual childcare structures and routines, with limited options for entertaining their children at home for prolonged periods of time.

> “He used to go to playgroup as well. So, every single day…we used to go out and do something…That just all stopped, so I had to find a way to make that work indoors at home just in our four walls” (ID337_couple_mum)

For some, the loss of usual support, with children not being at school or in childcare, affected their ability to work and subsequently generate household income, which in turn exacerbated stress.

> “I was planning on taking up shifts, but because lockdown happened, I then couldn’t pick up shifts because I had all four kids at home…my mental health just dropped dramatically when I realised I had no job, the kids were at home full time” (ID301_lone_mum)

Some participants who were self-employed or freelance workers, had some flexibility around the amount of time they spent looking after the household and children and taking on less work. In lone parent households with no other source of income, however, this reduction in work to look after the family led to worries about financial security and increased stress.

> “I’m having to face up to having difficult conversations, possibly move, can’t commit to things, can’t pay everything at the moment. It feels like a very strange situation to be in having been self-sufficient for so many decades.” (ID382_lone_mum)

#### Lack of control

Some participants described the inability to control what was happening to their home life, work and daily routines as difficult, but that the consequences may have a favourable effect on other areas of their life.

> “I realised that it’s possible to have no control because, yes, having my work paused and income paused, which again was helpful in some ways, because my children were home.” (ID382_lone_mum)

Participants also raised concerns about suddenly being unable to control access to essential items and whether they would be able to provide food for their family.

> “It was so weird that half the shelves were empty. It was just like what is going on? Where is this going to go? What is happening? This is the most traumatic change, nationally and globally, that I’ve experienced in a way. When my daughter said to me…why is there no food on the shelf? I really, genuinely had a moment of what if you can’t feed your kids?” (ID401_couple_mum)

Mothers in particular spoke of their difficulty coping with these feelings of uncertainty caused by a perceived lack of control. For some, this was frightening and overwhelming.

> “But it’s the not knowing, it’s the constant not knowing, and the constant changing. You’re never on firm ground, it’s always like quicksand. You sort of just stand on it and then something else happens.” (ID395_couple_mum)

### Changes to usual support networks

#### Impact of childcare/ education closures

Loss of childcare support came in many forms, including closure of nurseries and schools, withdrawal of home-based childcare and loss of informal care. Prior to the pandemic, many families relied on childcare support from the wider family, such as grandparents. However, with restrictions to household mixing and concerns about risks of COVID-19 to older people, this was no longer a viable childcare option for most. Instead, childcare responsibilities often shifted to mothers.

> “Well, really it’s all our parents that we would’ve relied on, obviously we couldn’t have that, so it was up to myself and my wife just to manage the kids. It was tough for her because I was going to work every day, I had that break from trying to home-school two children, and one of them having special educational needs, so it was really, really tough for her. I had the easy gig, being able to go to work.” (ID392_couple_dad)

In the absence of usual childcare and support structures, many participants described their fear of being unable to cope if both parents contracted the virus at the same time and how they would manage to look after their children.

> “One really big worry at the start was that my husband and I would both get COVID at the same time and given what we were hearing about just how unwell some people felt, even if they recovered within a few weeks, we were genuinely worried about our ability to cope with the children on our own.” (ID394_couple_mum)

Some families found forming a support bubble a solution. In one case, a parent and her young child moved in with the child’s grandparents.

> “But for me and my son, the fact that we’d moved in with my mum and dad. I think I would have really struggled if it was just me and him, and our house is quite small, so the fact that I have that support and I was lucky enough to be able to move in with them really helped.” (ID391_lone_mum)

#### Life moving online

For some participants with school-aged children, alternative support came from online resources, for example, school lessons or nursery sessions conducted remotely. Success varied depending on the quality of provision, the resources available at home, and the motivation, age, and independence of the children.

> “Home schooling for my daughter (secondary school) was fairly easy, because the school set up online lessons. Not face-to-face camera lessons, but just doing your work online, on Google Classrooms. That’s fine, and she’s self-motivated and she can get on with it. And she understands, and she can message her teachers, and she could message her friends as well. But for my (younger) son, it was a really hard…He effectively lost four months; I would say.” (ID405_couple_dad)

One participant described the stress imposed by digital exclusion, finding the shift to remote working and schooling extremely difficult due to limited internet access at home.

> “I don’t have Wi-Fi in my house. And so that made life difficult. The university paid me to get this, I don’t know what it’s called, a portable Wi-Fi. But the signal quality on it was just rubbish and it was just taking me so long to try and open up anything…So, I’d get to the end of the day and I’d feel tired and stressed and I probably would have literally achieved nothing…I haven’t done any work, I felt like I wasn’t teaching my son and it was awful. (ID386_lone_mum)

For parents with younger children, online alternatives to childcare were particularly unhelpful, sometimes becoming a source of stress as opposed to a solution.

> “So, we did do a Zoom online activity, which was horrible, because the point of the nursery is, he’s out of the house. And I have a three-month-old and I’m having to do this cutting-out stuff with him in front of a camera. And he just runs off and she cries. And I’m, like, what do you want me to do? Why am I even trying to attend this?” (ID422_couple_mum)

### Changes to personal relationships

Relationships within and beyond the immediate family were discussed by all participants. Maintaining positive relationships was important for parents’ wellbeing, however, participants also described both emotional and practical barriers that prevented this.

#### Strengthened family relationships

Many participants, particularly fathers, valued being at home and spending more time with their partner and children.

> “The positive stuff around being at home and getting to spend time with my son, and with my wife, that we otherwise wouldn’t have.” (ID381_couple_dad)

For some fathers, spending more time with their family led to better communication between family members and an enhanced understanding of their children’s needs.

> “So, we’re now beginning to sit down together as a family and say okay, this is what we need to do, so we’re planning very well. So, in a positive way, we now know why he does what he does” (ID297_couple_dad)

Being at home also enabled some participants to observe positive developments in sibling relationships that might not have occurred when older siblings were at nursery or school and parents were at work.

> “Looking at his relationship with his sister grow. She just adored having her brother home. And looking at him becoming more aware and empathetic of her was really positive.” (ID394_couple_mum)

#### Challenges to maintaining positive relationships

Participants with children of different ages described feeling guilty over difficulties in managing relationships between siblings, particularly given the limited opportunities for socialising with other children their own age.

> “One of the things that I felt really bad about was my youngest…because, obviously the age gap between the children, although my two youngest are close together and my two older are close together, my youngest son is very mature and very advanced for his age. So, he hangs out with the two oldest. So those three come together and they’re all quite happy. But of course, my daughter gets left out.” (ID301_lone_mum)

In families where there was one child, filling the void caused by a lack of interaction with their friends was a source of discomfort for some parents.

> “And he needs to see his friends because it’s difficult, he’s at that age where he wants to do imaginary play. And I don’t know whether it’s because I’m stressed out or it’s just me, but I can’t do that type of play. I can play a board game, I can play a computer game, but I can’t, I’m not a (primary school aged) boy. And I find that stressful…letting my son down.” (ID386_lone_mum)

Relationships with partners were the focus of fluctuating tensions. While many described valuing more time with their partner at home, the lack of time apart also became a source of disagreement. Participants described a variety of stressors; for one mother, financial pressure added to conflict with her partner.

> “He’s at home a lot and he also lost his job, and he can’t find a new one because of the pandemic… And he’s a full-time student now… So, there is always that financial pressure and, of course, it adds to the sources of our arguments, of who’s going to work and all these things.” (ID422_couple_mum)

#### Loss of physical contact

The sadness that participants felt about not being able to see their wider family, such as grandparents and cousins was described by many.

> “We found it really hard when we had to be separate from my sister and my nephew. Normally we do see them a lot and we’re really close with them…So, it was really weird not seeing him at all, and then just seeing him at the side of the road, or at the park, and not being able to go near him. So, that was challenging.” (ID391_lone_mum)

Online communication was commonly used to connect with the wider family, and although many were thankful for this option, others felt disappointment that face-to-face contact had been lost. Some felt saturated by work video calls and therefore did not benefit from communicating with extended family online.

> “My parents would Zoom my children fairly regularly, once a week. They didn’t Zoom before this. Telephone calls would be more common. I didn’t join in on the Zoom a lot. I was finding I was all virtual world done by the end of a workday, and really didn’t want to talk at all, to anybody, because I spend all my day talking.” (ID384_lone_mum)

For parents whose social networks were geographically close by, and who usually socialised with friends and family face-to-face, the shift to remote communication was experienced as unfamiliar, undesirable and led to increased social isolation.

> “That face-to-face is not there, and that’s what I really, really miss the most. I don’t like this aspect of talking to somebody on the phone, in fact, at times, I don’t even answer my phone, that’s something, again, I don’t do” (ID297_couple_dad)

Beyond family, the support of friends was essential to most participants for maintaining positive mental health and wellbeing, although many also missed physical contact with close friends.

> “I think… messaging people on WhatsApp who might be having a crisis, or feeling a bit low, it’s just not the same as seeing each other, and I struggle with that. I’d love to be able to go round to someone’s house, and have them come round to mine, and have that conversation face-to-face.” (ID414_couple_mum)

#### Colleagues as a source of support or stress

Having supportive and understanding colleagues helped to reduce the stress of managing work and home life concurrently; however, fathers were more likely to describe feeling well-supported.

> “I’m lucky where I work, where they do understand that parents need probably a bit more slack when it comes to working from home during the pandemic, and if I say, look, I need an hour just to do all my kids’ stuff, fine, take an hour. So, I’m quite lucky where I work, they’re very understanding of the work-life balance that’s required.” (ID399_couple_dad)

Conversely, mothers had fewer positive experiences with issues such as loss of income, unemployment, job insecurity and redundancy all adding to feelings of stress.

> “At that point, there was no childcare available and I got into a really difficult situation with my work who wanted me to come back, but I had no childcare. And so, they just point blank refused to furlough me.” (ID400_couple_mum)

### Strategies for coping

#### Protective activities

Participant wellbeing was at times affected from managing with children in the house for long periods of time, without the usual variety provided by friends, social activities, and going to work or school. To cope with these multiple challenges imposed by the pandemic, all participants cited using protective activities to maintain their wellbeing. Many participants discussed the importance of a spacious home environment and access to outdoor space, including private gardens or local community green spaces.

> “We didn’t leave the house…All exercise and everything happened indoors and like I said, thank god we have a garden, and we were able to break things up. We were able to make it fun, we were able to make it work…So, we did it until…the first lockdown restriction was lifted.” (ID337_couple_mum)

Equally, time and space away from family was important for most participants. For some, mainly mothers and lone parents, spending time outside of the home was essential for their wellbeing.

> “In the evening also, I’d go out to the shop for an hour…just to be alone or just to talk to a friend…because you need a space on your own to talk to a friend at the moment.” (ID422_couple_mum)

Some participants described how they managed the lack of structure and loss of routine imposed by the pandemic by implementing practical tools and creating new daily routines to help themselves and their family cope.

> “On a practical front, day three, I got my husband to buy a whiteboard from Amazon and our nice picture on the kitchen wall got binned and there was a whiteboard running tallies of what we were doing in the day, what we needed in shopping, what was coming up. All that kind of stuff. Just to try and externalise some of that and not have it in our heads all the time.” (ID394_couple_mum)

For other participants, accepting that events were out of their control contributed to positive wellbeing and opportunities to re-engage with enjoyable activities.

> “Life gives you lemons you make lemonade, or the glass is always half full in my eyes. You’ve just got to make do with what you’ve got really, and it was an opportunity for me to get back into the fitness that I so loved.” (ID337_couple_mum)

Though many participants recognised the value of the protective activities they were adopting or wished to adopt, having to manage multiple responsibilities within the home sometimes made participating in these activities difficult.

> “But it was very difficult to find the time and headspace in the day. And in some ways, it was quite frustrating, because it felt like I knew that there were all these resources available, and can I use mindfulness, and can I join a Zoom yoga class, and could I do all these things? And actually, there was just no time. You got up in the morning with the kids. When they went to bed, you cleaned the house, and had a shower and got ready for the next day. And so I feel like, in a different situation, I might have been much more resourceful in what I could do to attend to my mental health. But, for the most part, it was day-to-day survival, at least in the beginning.” (ID394_couple_mum)

For others a lack of motivation or fatigue brought on by constantly thinking about the pandemic and social distancing restrictions limited their efforts to maintain good mental health.

> “I think the exercise issue I’ve got to address. And I know I’ve got to address it, and find the time and space, but the mental exhaustion all day, and every day, thinking about COVID-related activities is…You want to do a bit of escapism, really.” (ID414_couple_mum)

For a small number of participants, their immediate environment was not conducive to engaging in protective activities, for example, living in cramped spaces with limited access to outside space and in close proximity to antisocial neighbours.

> “Where I’m at, at the moment, this is a one-bedroom flat. We have new tenants move across the street. In the evening at 11 o’clock, music is just playing so loud. What do we do? My wife says, let’s close the window. It is about 35 degrees, too hot. We don’t know what to do…So, all these little things just really, really crank me up. I’ve got to be lying there, I want to sleep, I can’t sleep down there because of the music.” (ID297_couple_dad)

#### Avoiding negative interactions and behaviours

In contrast to active participation in exercise and social activities, many participants described purposively avoiding certain activities to maintain their wellbeing. These included limiting contact with friends or family members perceived as unhelpful or in disagreement over the social distancing rules, as well as attempts to avoid triggering arguments in the home.

> “Now and again I’ll slip into that and I’m like, oh, I shouldn’t have done that because another can of worms has been opened. But I’ll try and avoid those difficult conversations. Because you’re in such close quarters with the people you live, you don’t have that break to then go away, digest, and physically just think about something different and then come back to a situation, it’s just all there, isn’t it? So, I’ve avoided those sorts of things.” (ID399_couple_dad)

Others described stopping or reducing their alcohol intake or being more mindful of limiting unhealthy eating behaviours.

> “I think try to keep eating healthily, so I guess you could phrase that as not to just vegetate and eat crap and loads of treats, which is easily done.” (ID381_couple_dad)

Another avoidance strategy described by some participants was to limit engagement with the rolling news coverage of the pandemic both on the television and social media.

> “Then I decided to go back to the news but only a couple of times a day and not look at all the scrolling news and updates. Just bite size. I’ve pretty much done that since and it’s helped a lot and it’s helped me put things into perspective a bit and focus on my here and now.” (ID401_couple_mum)

## Discussion

This study investigated the impact of the COVID-19 pandemic on the mental health and wellbeing of parents with young children in the UK. Findings revealed difficulties in navigating multiple responsibilities within the household and changes to previous home life structures and routines. Moreover, usual support networks and personal relationships were disrupted by the pandemic. Despite these challenges, our findings provide evidence of positive experiences stemming from lockdown measures, including accounts of strengthened family relationships. In attempts to cope with the multiple challenges to their mental health, most participants cited protective activities that they found helpful, including accessing outdoor space, spending time and space away from family, and avoiding unhelpful triggers.

Our findings align with existing research on the negative impact of the pandemic on the mental health and wellbeing of parents (19-21), however, our study helps to specifically explain why parental mental health was uniquely affected during this time. Having access to social networks and social support has been hypothesised to buffer the negative impact of stress (22, 23) and empirical work on the mental health consequences of natural disasters has identified the importance of social support as a protective factor against psychological stress (24). Previous research has also shown that in times of increased stress, people seek out familiar contacts rather than new connections (25). In line with these findings, participants in our study described reduced access to existing social support structures, coinciding with the need to navigate and juggle multiple responsibilities as a key source of stress, which was especially felt among mothers and lone parents.

The pandemic altered everyday life for many families, with participants in our study identifying disruption to their daily routines as a source of distress. Previous research has found that disruptions to regular routine and daily activities exacerbated the negative mental health impacts of exposure to psychologically stressful events (24). In our study, these disrupted routines arose when parents became responsible for home-schooling alongside their existing schedules and these pressures were felt most greatly when there were suggestions of conflicting priorities between parents. Notably, while previous research suggests that women took on more unpaid care work during the pandemic (13) leading to increased stress (26), some mothers in our study reported that this increased stress was rooted in perceptions that the pandemic had reinforced out-dated gender roles regarding parental responsibilities, with concerns that their children had noticed these stereotypes. It is also worth noting that some of the stressors experienced by parents existed prior to the pandemic, such as difficulties balancing childcare and work, and pressures to fulfil traditional gendered parenting roles (27).

For many participants in our study, particularly fathers and couples who continued to work from home throughout the pandemic, spending more time with their family and children was a positive experience. There is evidence that working parents will seek to retain the capacity to work flexibly, even after educational and caring arrangements are fully operational (28). Autonomy in future working patterns may alleviate the existing challenges parents have faced when navigating work life and family obligations (29), which may particularly benefit parents of children with disabilities or complex care needs (28). Changes in working practices post-pandemic may also go some way to reshaping the gendered dynamics of parenting. Although mothers have been more likely to forego paid work during the pandemic (either voluntarily or through temporary or permanent job loss) and have picked up a greater share of domestic responsibilities, there is also some evidence that fathers took on a greater share of childcare responsibilities than before (13). How parents allocated work and childcare duties during the pandemic could therefore set a precedent for a more permanent realignment of priorities post-pandemic. Workplace support strategies that enable parents to manage their work lives and childcare flexibly and effectively alongside their own priorities are therefore needed.

Many parents have identified worries about their children’s wellbeing as a common stressor throughout the pandemic (8, 30) and our study identified that pressures were felt differently depending on the number and age of children within the household. Parents with one child or with multiple children of varying ages found meeting their needs challenging in the absence of opportunities for social interactions with their peers. Conversely, families with siblings who were content playing with one another found the experience of self-isolating less demanding. While previous research has focused on parent-to-parent peer interventions for improving parent mental health (31), strategies and interventions that facilitate children to connect with their peers may also alleviate stress among parents. Although we found evidence of attempts to facilitate children’s peer-to-peer interactions online, for example, remote nursery sessions, this was often not a replacement for face-to-face interaction or care. However, given that some parents found remote connectivity valuable both for themselves and their children, this reinforces the importance of families being equipped with suitable technology to assist with their wellbeing during this pandemic and in future similar scenarios.

While we focused on parents’ individual experiences in this study, their coping and adaptive behaviours deployed in the wake of COVID-19 may have also been affected by social dynamics within the family network (32), which may be an avenue for future research to explore. In common with other groups of people who were interviewed during the pandemic (33-35), some parents said they were able to cope by adding structure and routine, prioritising a change of scenery during the day for the family, or receiving additional support from social contacts. Whilst it is encouraging to see that individuals engaged in such protective behaviours to maintain wellbeing, it is of equal importance to support the maintenance of ‘positive’ outcomes from the pandemic in the future. Relatedly, some parents actively avoided ‘trigger’ activities or behaviours to safeguard their mental health and wellbeing during the pandemic, including active avoidance of arguments and reduced engagement with pandemic related news. These findings are in line with research with other groups that identified associations between following COVID-19 related news stories and poorer mental health (14, 33, 35). Engaging in avoidant coping strategies may protect against psychological distress in the short term, however research suggests that behavioural disengagement and denial leads to poorer mental health in the longer term (36). Support for parents to help facilitate their own healthy coping behaviours, including increased access to mental health interventions (such as cognitive behavioural therapy) or stress-reducing techniques (such as physical activity), should therefore be actively promoted by mental health services and employers (37).

A strength of this work is that it is the first known in-depth qualitative study to explore the experiences of parents with young children living in the UK during the COVID-19 pandemic. However, there are several limitations that need to be considered when interpreting the findings. First, whilst the data set is comprised of a large qualitative sample that generated rich and detailed accounts of parental life during the pandemic, the majority of interviews were conducted in the Autumn of 2020. This is a period when most children in the UK had returned to school. Although parents reflected on their own experiences of the pandemic as a whole and recalled that periods of home schooling added to stress, for many, the time at which interviews were carried out may well have been a time of lower stress and comparatively improved resilience. Second, the sample may be biased toward those parents motivated or willing/able to participate, and we are particularly missing the views of those who lacked access to the internet. Finally, our sample was highly educated and predominantly consisted of mothers, however where pertinent to do so, we have highlighted the views and differences in experiences between these groups.

## Conclusions

In summary, our study highlights the unique stressors experienced by parents of young children living in the UK during the COVID-19 pandemic. The findings suggest that employers must be cognisant of the challenges the COVID-19 pandemic has placed on parents of young children, particularly mothers and lone parents, and consider how appropriate flexible working arrangements, access to technology and mental health and social support would relieve stress and increase productivity. It is also essential that adaptive rather than avoidant coping strategies are harnessed and encouraged by employers and policymakers to promote the positive wellbeing of parents during times of stress both during the pandemic and as we transition into potentially new ways of working going forward.

## Data Availability

The data are not publicly available due to their containing information that could compromise the privacy of research participants.

## List of abbreviations

COVID-19: Coronavirus disease 2019
UCL: University College London
UK: United Kingdom
UKRI: United Kingdom Research and Innovation

## Ethics approval and consent to participate

Ethical approval to conduct the study was granted by the University College London research ethics committee (Project ID: 14895/005). All participants provided written informed consent to take part in the study.

## Availability of data and materials

The data are not publicly available due to their containing information that could compromise the privacy of research participants. The interview topic guide is available in Additional File 1

## Authors’ contributions

DF conceived the study and DF & AB designed the study. JD, AM, & AB collected data for the study, analysed and interpreted the data. JD wrote the first draft with input from TM. All authors provided critical revisions, read and approved the submitted manuscript. All authors had full access to the data in the study and can take responsibility for the integrity and accuracy of the data.

## Acknowledgements

The authors would like to thank Henry Aughterson and Katey Warran for their assistance during the recruitment and analysis stages of this paper. The authors would also like to thank those people who gave up their time to take part and contribute to the study.

